# Central oxidative stress and early vocational outcomes in first episode psychosis: A 7-Tesla Magnetic Resonance Spectroscopy study of glutathione

**DOI:** 10.1101/2021.09.17.21263506

**Authors:** Michael MacKinley, Sabrina D. Ford, Peter Jeon, Jean Théberge, Lena Palaniyappan

## Abstract

Following the first episode of psychosis, some patients develop poor social and occupational outcomes, while others display a pattern of preserved functioning. Several lines of evidence from preclinical, genetic and biochemical studies suggest a role for high oxidative stress in poor functional outcomes. The measurement of intracortical glutathione (GSH) using magnetic resonance spectroscopy (MRS) provides an opportunity to investigate the relationship between central antioxidant tone and functional outcomes at the time of first episode psychosis (FEP). A body of epidemiological studies indicates better functional outcomes in patients at early stages of schizophrenia compared to patients at a chronic, established phase of illness. We scanned 57 patients with FEP and 30 matched healthy controls and estimated GSH resonance using 7-Tesla MRS. We minimised the confounding effects of illness chronicity, long-term treatment exposure and metabolic complications by recruiting patients with <2 weeks of lifetime antipsychotic exposure on average and followed up this cohort for the next 1 year to determine functional outcomes. Patients with FEP who achieved employment/education or training status (EET) in the first year, had higher GSH at the baseline than healthy controls. Social and occupational functioning assessment scale (SOFAS) scores were also significantly higher in patients with higher GSH levels at the outset, after adjusting for various confounds including baseline SOFAS. Patients who were not in employment, education or training (NEET) did not differ from healthy subjects in their GSH levels. Our observations support a key role for the central antioxidant tone in the functional outcomes of early psychosis.

## INTRODUCTION

For patients with schizophrenia, the probability of functional recovery is highest at the early stages of the illness, around the time of the first psychotic episode^1–3^; when a chronic pattern of the illness gets established, the chances of functional recovery greatly diminish, with only a small sub-group (∼13%) recovering at this stage ^4,5^. Currently, we do not know what mechanistic processes underlie these diminishing returns in recovery rates over time. Several clinical characteristics (e.g., the presence of negative, disorganised symptoms, cognitive deficits^6^) have been observed in association with poor functional outcomes; in particular, the degree of functioning at first presentation (baseline or premorbid) explains a significant amount of variance in long term functional outcomes^7–9^. Despite their explanatory power, these clinical associations do not offer an actionable mechanistic marker that can be harnessed for therapeutic purposes. There is an urgent need to understand the neurobiological factors that contribute to differences in functional outcomes in early stages of illness.

One promising approach to study variable outcomes in psychosis is quantifying the relative burden of oxidative stress experienced by patients during the first psychotic episode^10^. Fournier and colleagues utilised a data-driven stratification procedure on a cohort of patients with early psychosis and identified a subgroup with superior functional outcomes; this subgroup was characterised by lower levels of oxidative stress markers (especially glutathione peroxidase) in the blood^11^. Lower baseline blood levels of glutathione (GSH) predict later cognitive deficits^12^ as well as brain volume loss in early psychosis^13^. While peripheral markers of oxidative stress correlate with concentration of the primary intracortical antioxidant glutathione^14^, a direct link between central glutathione levels and functional outcome in first episode psychosis (FEP) is yet to be demonstrated. Wood and colleagues^15^ reported a 22% increase in medial temporal GSH levels in first episode psychosis; in a sub-sample from this study, treatment related increase in GSH was associated with a gain in global functioning scores^16^. In a small group of individuals with various mental health difficulties indicating a high-risk state for psychosis, we recently demonstrated higher GSH levels measured using magnetic resonance spectroscopy (MRS) from the dorsal anterior cingulate cortex (ACC) in those with better social and occupational functioning^17^. In the current study, we investigate if ACC GSH levels measured at the onset of psychosis, before establishing regular antipsychotic treatment, predict later functional outcomes in the first year of early intervention.

Early functional outcome status is a well-established indicator of long-term course of schizophrenia^18,19^. An exciting translational possibility of linking GSH levels with functional outcomes in psychosis, is the availability of targeted treatments that can improve intracortical GSH in patients. Several clinical trials have reported on the safety and efficacy of the glutathione precursor N-acetylcysteine (NAC) in patients with psychosis^20^. These trials (6 RCTs)^21^ indicate that NAC produces a modest, but significant improvement in cognitive deficits and negative symptoms (critical determinants of poor functional outcomes), when used as an adjunct to antipsychotics. Thus, in patients with psychosis, a deficit in intracortical GSH is likely to be a potentially modifiable pathway of poor outcomes.

## METHODS

### Participants

The sample for the present analysis consisted of 72 new referrals to the PEPP (Prevention and Early Intervention for Psychosis Program) at London Health Sciences Center. After exclusions were made due to missing/poor quality scan data our final sample consisted of 57 patients (48 males/9 females) (Table 1). All participants provided written, informed consent prior to participation as per approval provided by the Western University Health Sciences Research Ethics Board, London, Ontario. Inclusion criteria for study participation were as follows: individuals experiencing first episode psychosis, with not more than 14 days of cumulative lifetime antipsychotic exposure, no major head injuries (leading to a significant period of unconsciousness or seizures), or known neurological disorders, and no concurrent substance use disorder.

**Table 1:** Demographic and Clinical Characteristics of Healthy Controls and Patients

| Variable | Healthy Control<br>n=30 | All Patients<br>n=57 | EET Patients<br>n=31 | NEET Patients<br>n=26 |
| --- | --- | --- | --- | --- |
| **Demographic Data** |  |  |  |  |
| Gender (Male/Female) | 19/11 | 48/9 | 25/6 | 23/3 |
| Age [ M (SD) ] | 21.57 (3.45) | 22.75 (4.28) | 22.16 (3.92) | 23.46 (5.29) |
| NS-SEC [M (SD) ] | 3.20 (1.42) | 3.34 (1.27) | 3.31 (1.11) | 3.42 (1.38) |
| Avg Months to NEET assessment |  | 9.00 (4.38) | 8.90 (3.96) | 10.04 (4.79) |
| **Clinical Data** |  |  |  |  |
| DUP [M (SD)] | N/A | 7.75 | 6.39 (7.02) | 5.79 (6.70) |
| Antipsychotic Defined Daily Dose Equivalents | N/A | 1.80 (2.81) | 1.62 (3.03) | 1.79 (2.09) |
| Baseline SOFAS [M (SD)] | 82.92 (4.20) | 41.04 (12.48)** | 43.94 (15.10)** | 36.85 (8.69)** |
| PANSS-8 Total [M (SD)] | 8.0 (0.00) | 24.98 (5.76)** | 24.07 (4.95)** | 26.13 (6.57)** |
| PANSS-8 Positive [M (SD) ] | 3.0 (0.00) | 12.35 (2.32)** | 12.03 (2.44)** | 12.75 (2.15)** |
| PANSS-8 Negative [M (SD) ] | 3.0 (0.00) | 7.22 (4.17)** | 6.63 (3.76)** | 7.96 (4.61)** |
| CGI- Severity | 1.00 | 5.19 (0.97)** | 5.10 (1.13)** | 5.29 (0.75)** |

M, Mean; SD, standard deviation; NS-SEC, national statistics socio-economic classification; DUP, Duration of untreated Psychosis in Months; NEET, Not engaged in employment education or training; EET, Engaged in employment education or training; SOFAS, Social and Occupational Functioning Assessment Score; PANSS-8, Positive and Negative Syndrome Scale – 8 Item Scale; PANSS-8 Positive, Positive and Negative Syndrome Scale total score for positive symptom items; PANSS-8 Negative, Positive and Negative Syndrome Scale total score for negative symptom items; CGI-Severity, Clinical Global Impression – Severity; \*Significantly different compared to healthy controls ( $p < 0.05$ ); \*\*Significantly different compared to healthy controls ( $p < 0.01$ ).

The mean lifetime total defined daily dose days (DDD × days on medication) for antipsychotic use was 1.80 days with 27 patients (47.4%) being completely antipsychotic naive at the time of scanning. Of those who had started antipsychotic treatment, (N=30; 52.6%), the median total defined daily dose days was 2.81 days (range of 0.4–14 DDD days). Patient consensus diagnosis was established using the best estimate procedure described by Leckman et al.^22^ following 6 months of treatment. Diagnoses were made based on the Structured Clinical Interview for DSM-5.

Healthy control subjects (n=30) were recruited through posters and word of mouth advertising. Healthy control subjects had no personal history of mental illness, no current use of medications, and no first-degree relatives with a history of psychotic disorders. Healthy controls were group matched to the FEP cohort for age and parental socio-economic status (the National Statistics Socioeconomic Classification: five-class version^23^). Similar to their FEP counterparts, those with a history of substance use disorders in the past 12 months, significant head injury or neurological disorders were excluded.

### Clinical Measures

A clinical examination was conducted during the baseline assessment (the same day imaging was performed) by a research psychiatrist for patients or a trained rater for healthy controls. The clinical battery was used to assess patient symptom severity and to ensure that control subjects were free from current Axis I disorders and history of either psychotic illness or neurologic disorder. To assess symptoms of psychosis the Positive and Negative Syndrome Scale – 8 Item (PANSS-8) was used^24^. The PANSS-8 is an abbreviated version of the 30 Item PANSS clinical assessment of symptomology in schizophrenia and psychosis with acceptable internal consistency and highly correlated with the full PANSS^24^. Items are scored on a 1 (absent) to 7 (extreme) Likert type scale, assessing both positive (P1-delusions, P2 – Conceptual disorganization, & P3-hallucinations), and negative domains (N1-Blunted or flat affect, N4-passive social withdrawal, and N6-impoverishment of speech).

Additionally, the Social and Occupational Functioning Assessment Scale (SOFAS) was administered at baseline and follow up. The SOFAS is a single item measure of functioning scored between 1 (persistent inability to maintain minimum functioning without external support) and 100 (superior functioning in a wide range of activities). The SOFAS ratings of social and occupational functioning were made independent of symptom severity. In our study SOFAS was taken with consideration for current functioning (rather than highest level of functioning over the past year).

We obtained baseline SOFAS on the same day MRI data were acquired. The patients enrolled in the PEPP clinic were followed up over the next 12 months, and we ascertained their NEET status and follow-up SOFAS scores between 6 to 12 months. The functional assessment was based on multiple sources of information: patient interviews, information from the psychiatrist providing the clinical care, PEPP case managers and, where required, information from family members documented in clinical charts. Due to the need for multiple information sources, not all patients were assessed at the same time point after their illness onset, but the vocational (NEET) status of the cohort between the window of 6 to 12 months was captured, in addition to baseline and follow-up assessment of cross-sectional functioning using SOFAS. Patients were classified as NEET (vocationally inactive) if they were unemployed and not in any form of schooling/education for more than half of the time since the onset of treatment for psychosis. Individuals who were engaged in work or school for more than half of the duration of treatment were classified as EET (vocationally active). This definition considers a longer time frame (up to 6 months) than the 1-week period used by the Organisation for Economic Co-Operation and Development (OECD^25^), in line with its use in early intervention services for psychosis^26,27^. A consensus was reached within the research study team when discrepancies noted in reported functioning between patient’s accounts and those of care providers.

### MRS assessment

A complete description of the MRS protocol used in the present study has been described in previous work by Dempster and colleagues^28^. Metabolite concentrations (glutamate and GSH) were estimated using single-voxel ^1^H-MRS data acquired with a Siemens MAGNETOM 7.0T head-only MRI scanner (Siemens, Erlangen, Germany) using an 8-channel transmit/32-channel receive head coil at the Center for Functional and Metabolic Mapping of Western University in London, Ontario. A 2.0 × 2.0 × 2.0 cm (8 cm^3^) voxel was placed on the bilateral dorsal ACC using a two-dimensional anatomical images acquired in the sagittal direction (37 slices, TR = 8000 ms, TE = 70 ms, flip-angle (α) = 120°, thickness = 3.5 mm, field of view = 240 × 191 mm). The posterior end of the voxel was set to coincide with the precentral gyrus and the caudal face of the voxel coincided with the most caudal location not part of the corpus callosum. The angulation of the voxel was determined to be tangential to the corpus callosum (Figure 1).

**Figure 1:**
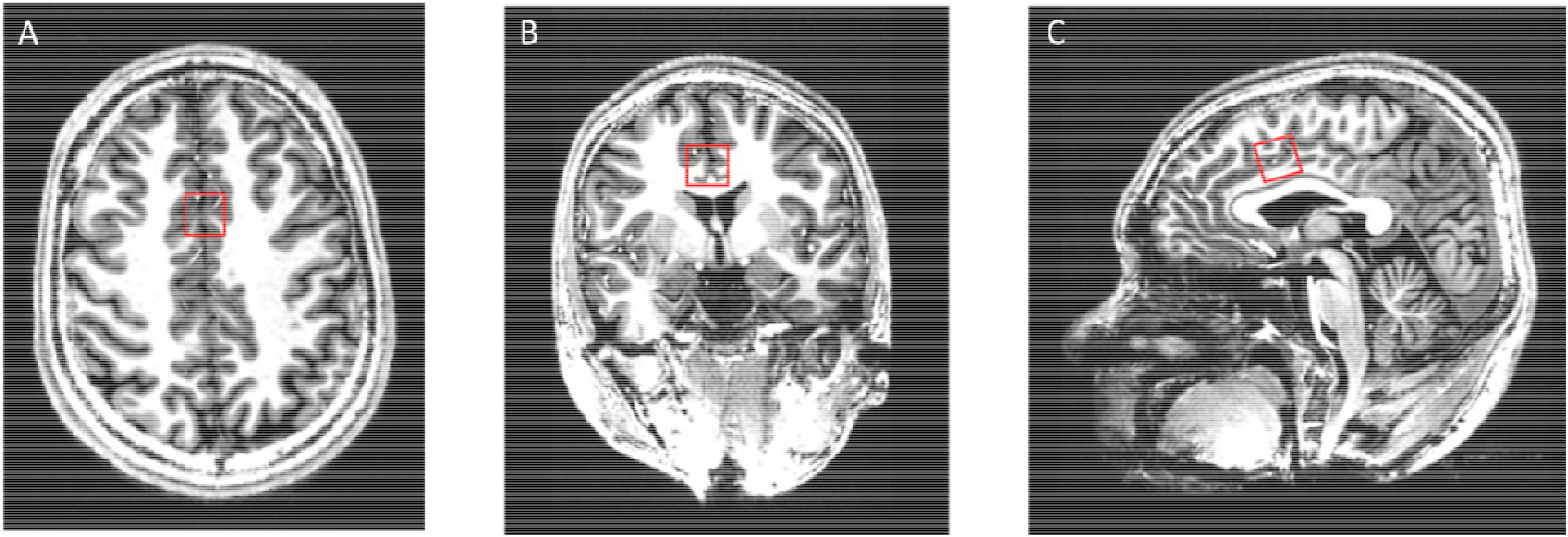
Illustrative example of the voxel placement on the dorsal ACC. A) Dorsal Brain View B) Posterior view C) Sagittal view

### Statistical analyses

We used IBM SPSS Statistics version 24 for all analyses. First, the primary hypothesis of a relationship between NEET status in the first year and glutathione was tested using *a one-way ANOVA*, with healthy controls, NEET and EET patients as 3 groups of interest. Second, within the patient group, a bivariate correlation between follow-up SOFAS scores and GSH measurement was conducted, with bootstrapping for generating p-values and confidence intervals. We also tested if GSH levels retain their ability to predict follow-up SOFAS, after adjusting for the variance explained by baseline functioning (SOFAS at the time of MRS scanning), age and gender as covariates. Finally, we excluded patients without a clear diagnosis of schizophrenia by 6-12 months assessment and tested the relationship between GSH and SOFAS and NEET.

## Results

When comparing healthy controls to FEPs, no statistically significant differences existed for age, parental socioeconomic status, or gender, although male participants were overrepresented in both groups (Table 1). As expected, there were several group-level differences in clinical scores between patient groups and healthy controls (Table 1). No significant differences in baseline demographic or clinical differences were identified between EET and NEET patients, with the exception of baseline SOFAS scores, which were higher among EET patients (t= 2.23, p= 0.031). Including subjects without useful MRS data did not affect this demographic profile (Supplementary Table 1).

### Group differences in GSH levels

Glutathione levels were significantly different among the 3 groups (FEP-NEET, FEP-EET and HC) at the p<0.05 level [F(2,84)= 4.55, p= 0.01]. Post hoc comparisons using the Sidak test indicated that the mean GSH levels for FEP-EET subjects was significantly higher than healthy controls [[Mean(SD) of GSH: FEP-EET= 1.76(0.37), HC= 1.49(0.32); p=0.01]. However, FEP-NEET subjects [M(SD) =1.60(0.34)] did not significantly differ from the healthy controls (p= 0.25) or FEP-EET (p= 0.57). Of note, patients as a whole had higher GSH levels than healthy controls (FEP= 1.68 (0.36), HC= 1.49(0.32); t(85)= 2.46, p= 0.016). These results are shown in Figure 2.

**Figure 2:**
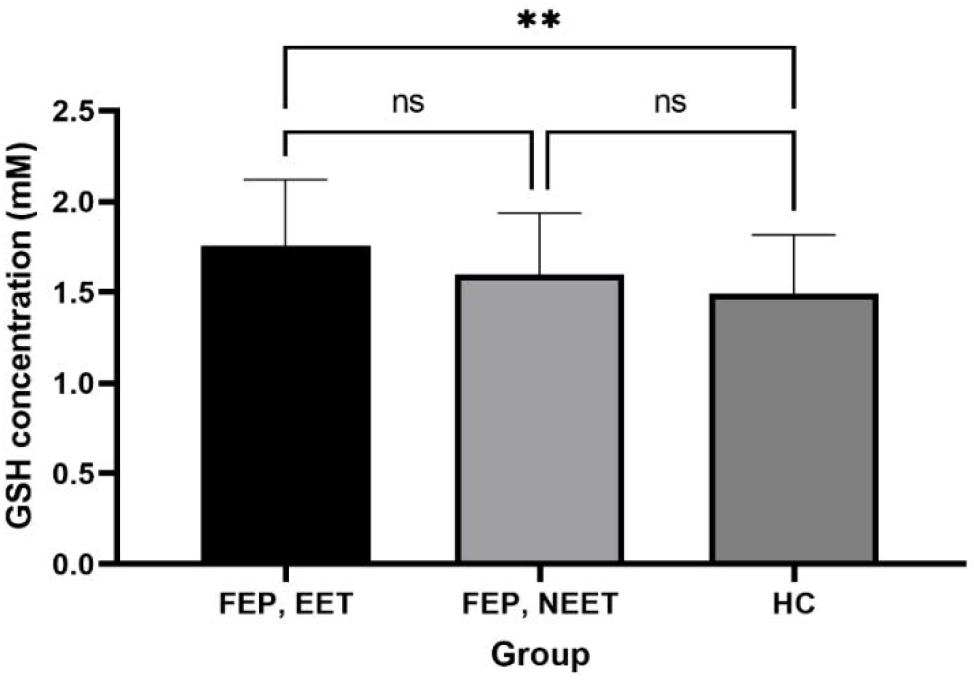
Relationship between GSH levels and SOFAS among healthy controls, EET and NEET patients

Within the FEP group, GSH measure at baseline was positively correlated with follow-up SOFAS scores r(44) = 0.41 [bootstrap 95% CI: 0.2-0.61], p=0.006. GSH levels, in combination with age, gender and baseline SOFAS scores, predicted follow-up SOFAS (R^2^= 0.29, F(4,36)= 3.76, p= 0.013). GSH levels (β= 0.36, p= 0.016) showed the strongest association out of all independent predictors for follow-up SOFAS, after adjusting for the variance explained by baseline functioning, age and gender (β= 0.17-0.21, all p> 0.15). The correlation between adjusted GSH levels and follow-up SOFAS is displayed, stratified by NEET status, in Figure 3.

**Figure 3:**
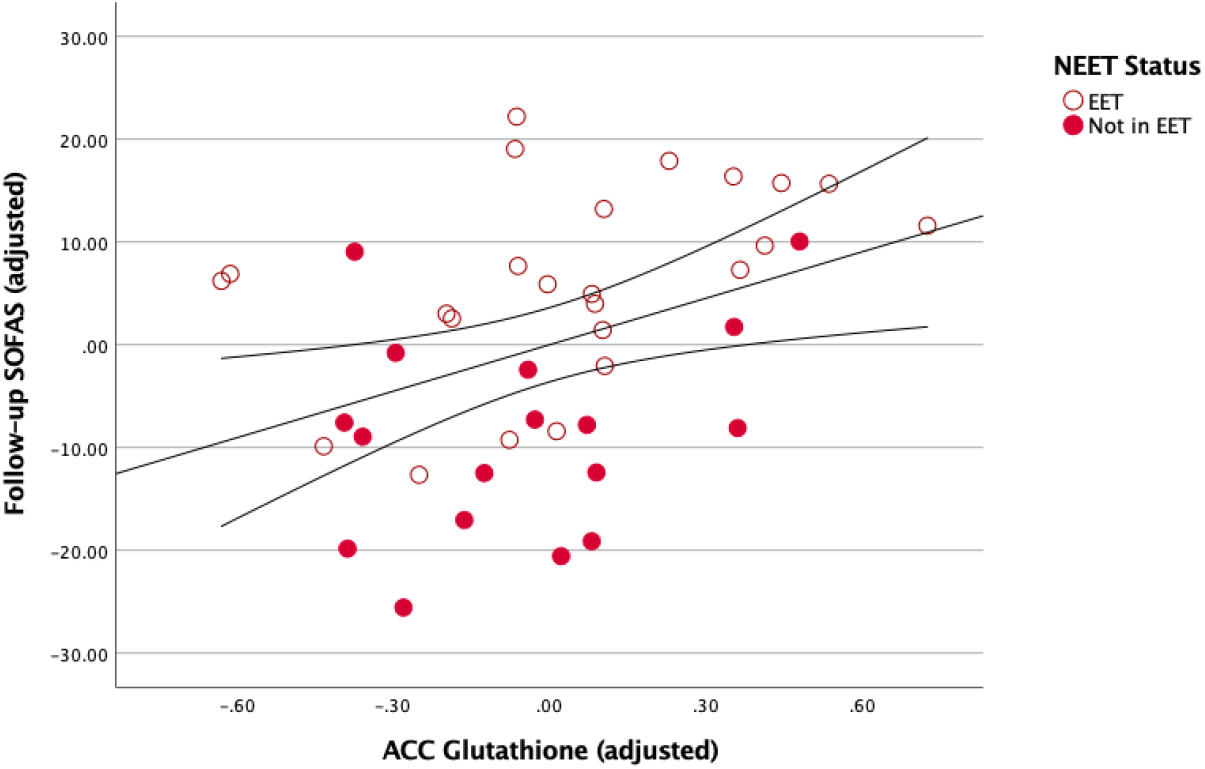

### Analysis restricted to first episode schizophrenia only

Finally, we repeated the above analysis in a subset of patients with a consensus diagnosis of schizophrenia/schizoaffective disorder by 6-12 months, after excluding 5 subjects with major depressive disorder, schizophreniform or bipolar disorder. Glutathione levels continued to differ significantly among the 3 groups (FES-NEET, FES-EET and HC) at the p<0.05 level [F(2,79)= 4.16, p= 0.02]. Post hoc comparisons using the Sidak test indicated that the mean GSH levels for FES-EET subjects was significantly higher than healthy controls [[Mean(SD) of GSH in FES-EET = 1.75(0.36), HC= 1.49(0.32); p= 0.016]. However, FES-NEET subjects [M(SD) = 1.59(0.35)] did not significantly differ from the healthy controls (p= 0.26) or FES-EET (p= 0.67).

Within the FES group, GSH measure at baseline was positively correlated with follow-up SOFAS scores r(39)= 0.38 [bootstrap 95% CI: 0.15-0.61], p= 0.017. GSH levels, in combination with age, gender and baseline SOFAS scores, continued to predict follow-up SOFAS in the FES group at a trend level, (R^2^= 0.24, F(4,32)= 2.47, p= 0.06). GSH level (β= 0.32, p= 0.05) continued to be the most prominent independent predictor of the follow-up SOFAS, after adjusting for the variance explained by baseline functioning, age and gender (β= 0.10-0.23, all p> 0.16).

### Prognostic relevance based on binarized GSH levels

To provide clinically relevant information to a patient with FEP whose GSH values have been quantified as described in this study, we performed a median split analysis of the patient group based on ACC GSH values of the entire sample (median= 1.586mM). We then compared the low-GSH and high-GSH FEP groups on the clinically relevant variables of NEET status and SOFAS at follow-up. The proportion of patients with FEP who were vocationally active (i.e., EET) significantly differed based on their baseline GSH status, with high-GSH FEP reporting 68% (23 of 34) in EET, while low-GSH FEP reporting 35% (8 of 23) in EET (Fisher’s exact p= 0.018). This translated to an odds ratio of 3.92 (95%CI: 1.34-11.2), and a relative risk of 1.95 (95%CI: 1.13-3.7) in favour of being in employment, education or training if an individual with FEP belonged to the high, instead of low-GSH group at the outset. The mean follow-up SOFAS scores for high-GSH FEP was also significantly higher than low-GSH FEP [Mean(SD) of SOFAS: high-GSH= 62.7(12.9), low-GSH= 53.5 (13.4); t (42)= 2.24; p= 0.03].

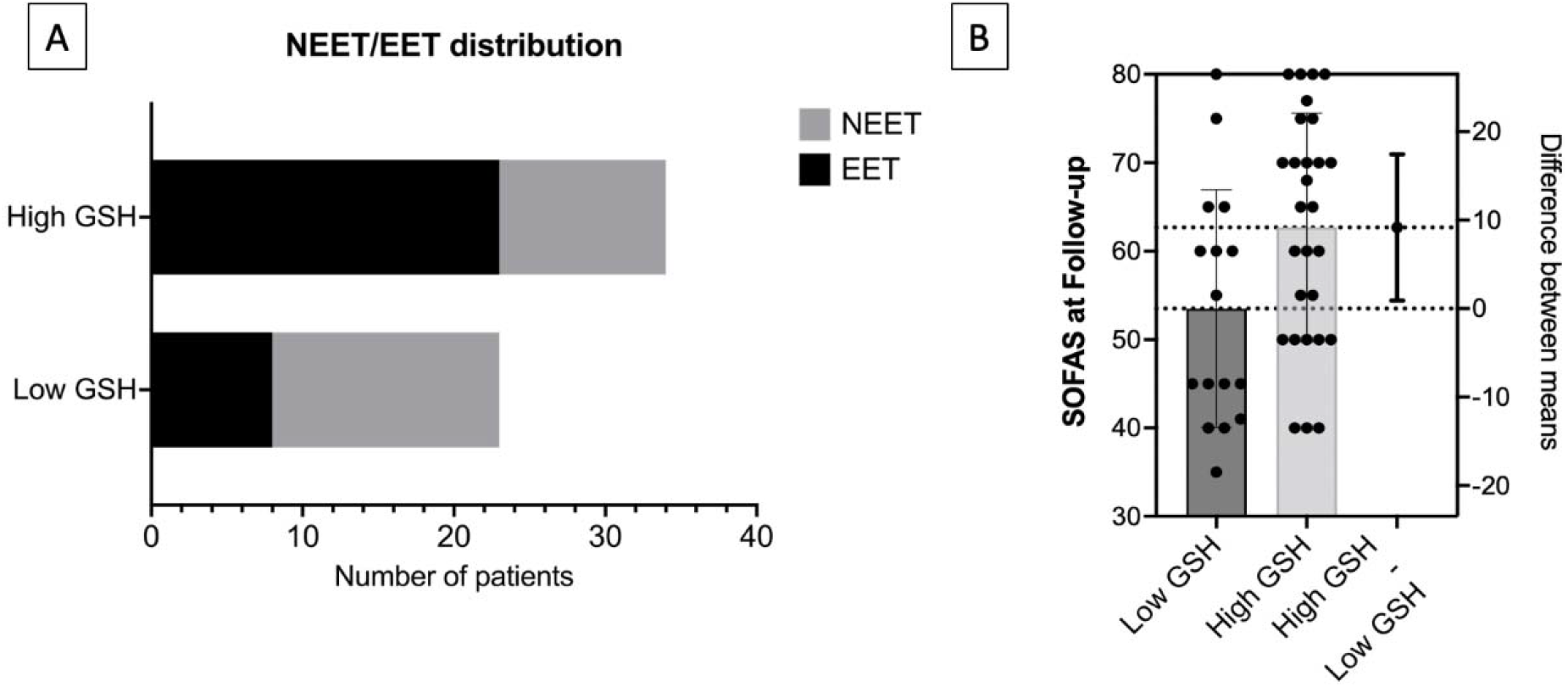

## DISCUSSION

Using 7T MRS to measure GSH concentration from ACC in untreated FEP subjects followed up over 1 year period, we report 2 major findings: (1) GSH levels are higher in patients with FEP when compared to healthy controls; this difference was driven by those who later displayed superior vocational status and socio-occupational functioning and (2) the baseline GSH levels predict later socio-occupational functioning over and above what can be predicted from baseline functioning, indicating a specific mechanistic role for the central antioxidant status in the outcome trajectories following first psychotic episode. Patients with GSH levels that were lower than the median values in the sample were at approximately two times higher risk of being vocationally inactive, with approximately 10 points lower scores in SOFAS.

Our observed relationship between lower GSH levels and lower SOFAS scores and NEET status is in line with several prior reports. In an overlapping sample, we previously reported a predictive relationship between low GSH and delayed response to antipsychotics^28^. Lack of early response is a critical indicator of long-term poor outcomes in schizophrenia^29–31^. In line with studies linking lower GSH to higher residual symptom burden^32^, negative symptoms^33^ and cognitive deficits^34^ in schizophrenia, our observation highlights a prominent ‘pathoplastic’ role for antioxidant status in shaping the outcomes of this illness. In fact, a recent 7T-MRS study of the ACC reported that patients who fail first-line antipsychotic treatments are more likely to have an intracortical GSH-deficit^35^. In our study, the presence of higher ACC GSH in FEP may indicate the overall superior treatment responsiveness in this cohort, given their untreated status. Such higher GSH levels have been previously reported in other brain regions in FEP^15,16^. The small to medium effect size difference also underscores the considerable heterogeneity in intracortical GSH levels in psychosis, in line with several 3T MRS studies that did not report a notable group difference in the ACC^36^ as well as other regions^37,38^. This heterogeneity may relate to individual differences (i.e., subgroups of patients with high or low GSH levels) or stage-specific differences (acute excess, followed by later deficit). While several exogenous factors may also affect antioxidant levels, there is no evidence for a major role for these confounders in explaining aberrations in glutathione pathway in psychosis^39^.

An important strength of our study is its longitudinal nature; with the temporal information we can establish that higher GSH levels at the outset predict and thus influence superior socio-occupational functioning seen over the next 1 year among patients. But a major limitation is the availability of a single time point of GSH levels. To establish a more conclusive causal inference, we need longitudinal follow-up studies that capture multiple time points from untreated early stages of psychosis to a stable phase when functional outcome trajectories become more established. Long term follow-up of experimental studies with patients in early stages of psychosis that receive adjunctive NAC will also be illuminating in this regard. We also lacked peripheral and genotyping measures of antioxidant capacity in this cohort; while these measures are more accessible than 7T-MRS, they do not consistently reflect intracortical GSH^40–42^.

We conclude that in patients with first episode psychosis in whom intracortical GSH levels are higher during the acute phase of psychosis, functional outcomes are superior to those with lower levels, over the next 1 year. Thus, ACC GSH measure may be a useful indicator of resilience to oxidative stress and functional recovery after first episode of psychosis. Taken together with prior MRS studies from established cases of schizophrenia indicating a profile of treatment failures and residual symptoms in patients with intracortical GSH-deficit, our observation makes a compelling case to investigate the role of pre-emptive antioxidant interventions in early stages of psychosis.

## Data Availability

The datasets generated during and/or analysed during the current study are available from the corresponding author on reasonable request.

## Acknowledgements

We thank Mr. Trevor Szekeres, Mr. Scott Charlton, Mr. Joseph Gati for their assistance in data acquisition and archiving. We thank all research team members of the NIMI lab and all the staff members of the PEPP London team, particularly Drs. Kara Dempster, Julie Richard, Priya Subramanian and Hooman Ganjavi for their assistance in patient recruitment and supporting clinical care. We gratefully acknowledge the participants and their family members for their contributions.

## Institutional Review Board Statement

This study was conducted according to the guidelines of the declaration of Helsinki and approved by the Research Ethics Board of the University of Western Ontario (Project #108268; October 19, 2020).

## Author Contributions

MM recruited patients, collected, and compiled demographic, functional outcome and treatment data and assisted in acquiring the scans. SF recruited patients, compiled demographic, functional outcome and treatment data and assisted in acquiring the scans. PJ acquired the MRS scans, performed the MRS quantification and analysis, supervised by JT. LP obtained the research funds, designed the project with JT and supervised MM, SF and PJ. LP conceived the analysis and wrote the first draft with MM and SF. All authors contributed to the writing and have approved the final version of the manuscript.

## Conflict of Interest

LP reports personal fees from Otsuka Canada, SPMM Course Limited, UK, Canadian Psychiatric Association; book royalties from Oxford University Press; investigator-initiated educational grants from Janssen Canada, Sunovion and Otsuka Canada outside the submitted work. All other authors report no relevant conflicts.

**Supplementary Table 1:** Demographic and Clinical Characteristics of Healthy Controls and Patients including those without useable MRS data

| Variable | Healthy Control<br>n=30 | All Patients<br>n=72 | EET Patients<br>n=37 | NEET Patients<br>n=35 |
| --- | --- | --- | --- | --- |
| **Demographic Data** |  |  |  |  |
| Gender (Male/Female) | 19/11 | 57/13 | 29/8 | 30/5 |
| Age [ M (SD) ] | 21.57 (3.45) | 22.97 (3.54) | 22.18 (3.81) | 23.47 (5.21) |
| NS-SEC[M (SD) ] | 3.20 (1.42) | 3.34 (1.27) | 3.31 (1.11) | 3.42 (1.38) |
| **Clinical Data** |  |  |  |  |
| DUP [M (SD)] | N/A | 7.75 | 6.39 (7.02) | 5.79 (6.70) |
| Antipsychotic Defined<br>Daily Dose Equivalents | N/A | 1.80 (2.81) | 1.62 (3.03) | 1.79 (2.09) |
| Baseline SOFAS [M<br>(SD)] | 82.92 (4.20) | 41.11 (12.96) | 43.94 (15.10) | 36.85 (8.69) |
| PANSS-8 Total [M (SD)] | 8.0 (0.00) | 25.28 (6.82) | 24.72 (6.56) | 26.39 (7.25) |
| PANSS-8 Positive [M<br>(SD) ] | 3.0 (0.00) | 12.39 (2.75) | 12.25 (2.76) | 12.93 (2.71) |
| PANSS-8 Negative [M<br>(SD) ] | 3.0 (0.00) | 7.38 (4.27) | 6.94 (4.07) | 7.96 (4.60) |
| CGI- Severity | 1.00 | 5.25 (0.98) | 5.16 (1.14) | 5.36 (0.87) |

M, Mean; SD, standard deviation; NS-SEC, national statistics socio-economic classification; DUP, Duration of untreated Psychosis in Months; NEET, Not engaged in employment education or training; EET, Engaged in employment education or training; SOFAS, Social and Occupational Functioning Assessment Score; PANSS-8, Positive and Negative Syndrome Scale – 8 Item Scale; PANSS-8 Positive, Positive and Negative Syndrome Scale total score for positive symptom items; PANSS-8 Negative, Positive and Negative Syndrome Scale total score for negative symptom items; CGI-Severity, Clinical Global Impression – Severity; \*Significantly different compared to healthy controls ( $p < 0.05$ ); \*\*Significantly different compared to healthy controls ( $p < 0.01$ ).

## References

1. Henry, L. P. et al. The EPPIC follow-up study of first-episode psychosis: longer-term clinical and functional outcome 7 years after index admission. J. Clin. Psychiatry 71, 716–728 (2010).

2. Addington, J. The promise of early intervention. Early Interv. Psychiatry 1, 294–307 (2007).

3. Dama, M. et al. Short duration of untreated psychosis enhances negative symptom remission in extended early intervention service for psychosis. Acta Psychiatr. Scand. 140, 65–76 (2019).

4. Jääskeläinen, E. et al. A Systematic Review and Meta-Analysis of Recovery in Schizophrenia. Schizophr. Bull. 39, 1296–1306 (2013).

5. Norman, R. M. G., MacDougall, A., Manchanda, R. & Harricharan, R. An examination of components of recovery after five years of treatment in an early intervention program for psychosis. Schizophr. Res. 195, 469–474 (2018).

6. Santesteban-Echarri, O. et al. Predictors of functional recovery in first-episode psychosis: A systematic review and meta-analysis of longitudinal studies. Clin. Psychol. Rev. 58, 59–75 (2017).

7. Díaz-Caneja, C. M. et al. Predictors of outcome in early-onset psychosis: a systematic review. Npj Schizophr. 1, 14005 (2015).

8. O’Keeffe, D. et al. The iHOPE-20 study: Relationships between and prospective predictors of remission, clinical recovery, personal recovery and resilience 20 years on from a first episode psychosis. Aust. N. Z. J. Psychiatry 53, 1080–1092 (2019).

9. Velthorst, E. et al. The 20-Year Longitudinal Trajectories of Social Functioning in Individuals With Psychotic Disorders. Am. J. Psychiatry 174, 1075–1085 (2017).

10. Murray, A. J., Rogers, J. C., Katshu, M. Z. U. H., Liddle, P. F. & Upthegrove, R. Oxidative Stress and the Pathophysiology and Symptom Profile of Schizophrenia Spectrum Disorders. Front. Psychiatry 12, 1235 (2021).

11. Fournier, M. et al. Implication of the glutamate–cystine antiporter xCT in schizophrenia cases linked to impaired GSH synthesis. Npj Schizophr. 3, 1–7 (2017).

12. Martínez-Cengotitabengoa, M. et al. Basal low antioxidant capacity correlates with cognitive deficits in early onset psychosis. A 2-year follow-up study. Schizophr. Res. 156, 23–29 (2014).

13. Fraguas, D. et al. Decreased glutathione levels predict loss of brain volume in children and adolescents with first-episode psychosis in a two-year longitudinal study. Schizophr. Res. 137, 58–65 (2012).

14. Xin, L. et al. Genetic Polymorphism Associated Prefrontal Glutathione and Its Coupling With Brain Glutamate and Peripheral Redox Status in Early Psychosis. Schizophr. Bull. 42, 1185–1196 (2016).

15. Wood, S. J. et al. Medial temporal lobe glutathione concentration in first episode psychosis: a 1H-MRS investigation. Neurobiol. Dis. 33, 354–357 (2009).

16. Berger, G. E. et al. Ethyl-eicosapentaenoic acid in first-episode psychosis. A 1H-MRS study. Neuropsychopharmacol. Off. Publ. Am. Coll. Neuropsychopharmacol. 33, 2467–2473 (2008).

17. Jeon, P. et al. Glutathione as a molecular marker of functional impairment in patients with at-risk mental state: 7-Tesla 1H-MRS study. 2020.11.17.20233635 https://www.medrxiv.org/content/10.1101/2020.11.17.20233635v1 (2020) doi:10.1101/2020.11.17.20233635.

18. Harrison, G., Croudace, T., Mason, P., Glazebrook, C. & Medley, I. Predicting the long-term outcome of schizophrenia. Psychol. Med. 26, 697–705 (1996).

19. Harrison, G. et al. Recovery from psychotic illness: a 15- and 25-year international follow-up study. Br. J. Psychiatry J. Ment. Sci. 178, 506–517 (2001).

20. Klauser, P. et al. T52. N-ACETYL-CYSTEINE ADD-ON TREATMENT LEADS TO AN IMPROVEMENT OF FORNIX WHITE MATTER INTEGRITY IN EARLY PSYCHOSIS. Schizophr. Bull. 44, S133–S134 (2018).

21. Yolland, C. O. et al. Meta-analysis of randomised controlled trials with N -acetylcysteine in the treatment of schizophrenia. Aust. N. Z. J. Psychiatry 54, 453–466 (2020).

22. Leckman, J. F., Sholomskas, D., Thompson, D., Belanger, A. & Weissman, M. M. Best Estimate of Lifetime Psychiatric Diagnosis: A Methodological Study. Arch. Gen. Psychiatry 39, 879–883 (1982).

23. Rose, D. & Pevalin, D. J. A Researcher’s Guide to the National Statistics Socio-economic Classification. (Sage Publications, 2003).

24. Lin, C.-H. et al. Early improvement in PANSS-30, PANSS-8, and PANSS-6 scores predicts ultimate response and remission during acute treatment of schizophrenia. Acta Psychiatr. Scand. 137, 98–108 (2018).

25. Youth and the labour market - Youth not in employment, education or training (NEET) - OECD Data. theOECD http://data.oecd.org/youthinac/youth-not-in-employment-education-or-training-neet.htm.

26. Iyer, S. et al. A NEET distinction: youths not in employment, education or training follow different pathways to illness and care in psychosis. Soc. Psychiatry Psychiatr. Epidemiol. 53, 1401–1411 (2018).

27. Maraj, A. et al. Caught in the ‘NEET Trap’: The Intersection Between Vocational Inactivity and Disengagement From an Early Intervention Service for Psychosis. Psychiatr. Serv. Wash. DC 70, 302–308 (2019).

28. Dempster, K. et al. Early treatment response in first episode psychosis: a 7-T magnetic resonance spectroscopic study of glutathione and glutamate. Mol. Psychiatry 25, 1640–1650 (2020).

29. Lambert, M. et al. Rates and predictors of remission and recovery during 3 years in 392 never-treated patients with schizophrenia. Acta Psychiatr. Scand. 118, 220–229 (2008).

30. Derks, E. M. et al. Antipsychotic drug treatment in first-episode psychosis: should patients be switched to a different antipsychotic drug after 2, 4, or 6 weeks of nonresponse? J. Clin. Psychopharmacol. 30, 176–180 (2010).

31. Carbon, M. & Correll, C. U. Clinical predictors of therapeutic response to antipsychotics in schizophrenia. Dialogues Clin. Neurosci. 16, 505–524 (2014).

32. Kumar, J. et al. Glutathione and glutamate in schizophrenia: a 7T MRS study. Mol. Psychiatry 25, 873–882 (2020).

33. Matsuzawa, D. et al. Negative Correlation between Brain Glutathione Level and Negative Symptoms in Schizophrenia: A 3T 1H-MRS Study. PLoS ONE 3, e1944 (2008).

34. Wang, A. M. et al. Assessing Brain Metabolism With 7-T Proton Magnetic Resonance Spectroscopy in Patients With First-Episode Psychosis. JAMA Psychiatry 76, 314–323 (2019).

35. Yang, K. et al. A multimodal study of a first episode psychosis cohort: potential markers of antipsychotic treatment resistance. 2021.05.03.442450 https://www.biorxiv.org/content/10.1101/2021.05.03.442450v1 (2021) doi:10.1101/2021.05.03.442450.

36. Das, T. K. et al. Antioxidant defense in schizophrenia and bipolar disorder: A meta-analysis of MRS studies of anterior cingulate glutathione. Prog. Neuropsychopharmacol. Biol. Psychiatry 91, 94–102 (2019).

37. Lesh, T. A. et al. Extracellular free water and glutathione in first-episode psychosis—a multimodal investigation of an inflammatory model for psychosis. Mol. Psychiatry 26, 761–771 (2021).

38. Iwata, Y. et al. Glutathione Levels and Glutathione-Glutamate Correlation in Patients With Treatment-Resistant Schizophrenia. Schizophr. Bull. Open 2, (2021).

39. Ballesteros, A. et al. No Evidence of Exogenous Origin for the Abnormal Glutathione Redox State in Schizophrenia. Schizophr. Res. 146, 184–189 (2013).

40. Gawryluk, J. W. Decreased levels of glutathione, the major brain antioxidant, in post-mortem prefrontal cortex from patients with psychiatric disorders. Int. J. Neuropsychopharmacol. 14, 123–130 (2011).

41. Zhang, Y., Catts, V. S. & Shannon Weickert, C. Lower antioxidant capacity in the prefrontal cortex of individuals with schizophrenia. Aust. N. Z. J. Psychiatry 0004867417728805 (2017) doi:10.1177/0004867417728805.

42. Coughlin, J. M. et al. A multimodal approach to studying the relationship between peripheral glutathione, brain glutamate, and cognition in health and in schizophrenia. Mol. Psychiatry 1–10 (2020) doi:10.1038/s41380-020-00901-5.

